# Developing an Individualized Clinical Prediction Rules of Antihypertensive Drugs: A Study Protocol Based on Real-world Practice

**DOI:** 10.1101/2020.01.29.20019364

**Authors:** Dongsheng Hong, Wendan Shi, Xiaoyang Lu, Wu Jiaying, Yan Lou, Lu Li

**Author notes:** is the corresponding author.

## Abstract

**Background:** Hypertension is one of the most urgent public health challenges, and drug therapy is the primary method to control blood pressure for patients. However, blood pressure control rate is still poor with antihypertensive drugs use. Although Clinical Prediction Rules (CPR) is useful to help clinicians make more appropriate decisions at the point of medication, the evidence is still limited in china. The objective of this study is to develop an CPR of antihypertensive drugs in individualized application of patients based on real-world practice.

**Methods:** A two-way cohort study has been conducted in one China’s large tertiary hospital using clinical information on patient characteristics, drug use and clinical outcome. Data extraction is through ICD-10 disease codes of hypertension from Electronic Medical Record System. Eligible patients admitted from September 2016 to August 2018 who have received at least one antihypertensive drug therapy is included. Patients were grouped into several exposure groups according to medications. COX regression model and clinical specialty survey is applied to identify Influencing Factors (IF) in different study groups, and the discriminant model was used to construct a CPR according IF. The accuracy of the CPR is analyzed by sensitivity, specificity, Youden’s index and Receiver Operating Characteristic (ROC) curve.

**Discussion:** Result is expected to provide valuable CPR for physicians and policymakers with respect to treating hypertension according characteristic of individual patients. By developing a predictive method for clinical outcomes and treatment costs of antihypertensive medication, we expect to discriminate those patients who would profit from specific scheme of antihypertensive drugs to minimal incidence probability of costs and complications in region of china.

**Trial registration:** This study was registered at www.chictr.org as a primary register of the WHO International Clinical Trials Registry Platform (ICTRP), and the registered number is ChiCTR1900026339.

**Highlights:** Although Clinical Prediction Rules (CPR) could recognize individual patient risk and help clinicians to make more appropriate decision at the point of medication as part of clinical decision support systems, the evidence in this respect is still limited in China.

This study is first going to construct the CPR of multiple antihypertensive drugs in real world practice of China.

The highlights of this study is aimed to provide a pragmatic method to support clinical decisions for patients who has received antihypertensive drugs before long-term diagnosis of hypertension in real world practice according to their characteristics that are accessible to clinicians.

## 1. Background

Hypertension is one of the constantly rising global health concerns^1, 2^, and it is considered as the enormous sponsor to the burden of disease^3^. In China, there are 270 million adults in total who suffered from a condition of hypertension, the Disability-Adjusted Life Year (DALY) has reached 37.94 million people per year^4^. The direct medical expenses for drug treatment are nearly 40 billion Chinese yuan per year, and it is still increasing accordingly^5^. Under such high medical expenditures, the control rate of Blood Pressure (BP) in China is only 16.9%^5^. With the aging population, changes in lifestyle, and population growth^6^, the burden of illness with hypertension is also growing over time.

Drug therapy is currently recognized as the primary method of hypertension treatment^7^. However, hypertension is normally an asymptomatic disease^8^. Patients may suffer from side effects of antihypertensive drugs (AHD), which bring about them become worse than they did before using medication. The poor personal response to standard treatment is one of the main reason of low rate of hypertension control^6^, and a large meta-analysis of prospective study stated the relationship between poor personal response (BP levels) and cardiac-cerebral vascular events^9^. To obtain the maximum antihypertensive effect of existing drugs, physicians and policymakers have turned to the rational use of antihypertensive drugs according to the characteristics of individual patient^10, 11^.

Although Clinical Decision Support Systems (CDSS) could be effective in providing an appropriate and cost-effective methods to improve the blood pressure control rate with reducing cardiovascular risk^12, 13^. The evidence of CDSS is from clinical guidelines, which major advocate evidence-based recommend and basically use a standard of universal treatment at the population levels^14^, are not highly concern to the characteristics of individual patients. Otherwise, AHD is more than 100 varieties in China that may have differences in terms of mechanism of action, dosage, adverse reactions and contraindications. The ability of an individual physician to optimize drug combinations for personal patient is rapidly going beyond their capabilities and comprehension^15^. Therefore, the clinical application of AHD should oblige to be person-centered for individual patients. It should balance of clinical effects, adverse reactions, and costs of treatment, in order to deduce which AHD will have the most effectiveness to control BP at an individual level with a good profile.

Clinical prediction rules (CPR) as an important component of CDSS, which is focusing particularly on the possible applications of individual characteristics for the evaluation risk of hypertensive patients. However, the evidence of CPR is still poor based on clinical feedback. Accordingly, the aim of this study is to eventually provide evidence of developing an CPR to identify of personal clinical characteristics that will better manage the AHD choice to improve disease control of the individual patient and overall hypertension population.

## 2. Methods

### 2.1 Study design

This research will conduct a two-way cohort study of hypertensive patients at the First Affiliated Hospital of College of Medicine of Zhejiang University in China (FHMZC), where the outpatient volume is moreover three million per year as China regional medical center. The information of Patient’s treatment outcomes will be collected prospectively in future, and the participants will be grouped according to their exposure at the time of recruitment. Exposure options are first-line treatment drugs according guidelines for hypertension in China^16, 17^, including Angiotensin-Converting Enzyme Inhibitor (ACEI), Angiotensin Receptor Blocker (ARB), Calcium Channel Blockers (CCB), diuretic, and β-blocker used as alone or in combination. This study will follow the statement of Strengthening the Reporting of Observational Studies in Epidemiology (STROBE), and The STROBE Statement Checklist of cohort studies is shown in online supplementary appendix 1.

The study period is planned to be over a 3-year period from 1 September 2016 to 31 August 2018. This period has been selected based on the availability of enough patients who would have been exposed to the study hypertensive drugs as well as coinciding with the availability of Electronic Medical Record System (EMRC).

### 2.2 Study cohort

The source patients include all EMRC patients aged 18 years and over, who has diagnosed with hypertension. Antihypertensive drugs exposure will be defined from dispensed prescription records of EMRC, beginning from 1 September 2016 and until the end of study. New patients using antihypertensive drugs will be determined by using a review observation period of 6 months prior to the index prescription date, and which must indicate no hypertensive drugs use during that period. The index date will be identified as the date of the new prescription for antihypertensive drugs. The group of study cohort would be constructed according to the antihypertensive drugs.

### 2.3 Data extraction

The study obtains data through data extraction of EMRC and follow-up survey. Data extraction will through ICD-10 disease codes^18^, including I10.X00, I10.X01, I10.X02, I10.X03, I10.X04, I10.X05, I10.X06, I11.900, I12.903 and I15.000. Information will be collected as the following: (1) demographic information; (2) medical history; (3) duration of illness; (4) underlying diseases; (5) medication history; (6) alcohol intake/smoking history; (7) inspection information; (8) efficacy evaluation; (9) complications.

### 2.4 Sample size

The sample size of our study is estimated by 10-15 times of the total factor^19^. If the factor involved is 30 temporarily, the sample size should be no less than 300 cases for a group. In addition, assuming that the loss rate of cohort study is 20%, the sample size should be further expanded to no less than 375 cases for a group. If the model is constructed according to 375 cases and verified by 375 cases, the final requirement is that the sample size should be enlarged to no less than 750 cases for a group.

### 2.5 Cost assignments and Outcomes measures

Direct costs will be assigned in Chinese Yuan (¥) using EMRC, and the costs is classified into two categories with drug related and costs for medical services. All major outcomes are identified: (1) coronary artery disease, (2) chronic heart failure, (3) renal insufficiency, and (4) cerebrovascular disease^20^.

### 2.6 Identification of influencing factors

Data characteristics among patients are commonly believed to be associated with treatment response and outcomes, and disease progression. We will conduct a method of COX regression and clinical specialty survey to identify Influencing Factors (IF) in different study groups of cohort^21^.

### 2.7 Individualized Clinical Prediction Rules

IF are directly related to medication in patients with hypertension. The discriminant model will be utilized to construct an individualized CPR based on the clinical outcomes and IF, and the clinical outcomes include the cost of treatment and the main outcomes as defined before^22^. The accuracy of the rules is analyzed by sensitivity, specificity, Youden’s index and Receiver Operating Characteristic (ROC) curve^23-25^.

The real-time data of each stage of patients are input into CPR to calculate the treatment cost of different medication schemes and their impact on the treatment outcome of patients under the current condition of exporting patients, so as to assist physicians in choosing Medication drugs for hypertension.

### 2.8 Statistical analysis

The measurement data of normal distribution are expressed by M (Q_R_), the classification data is worked out by relative number, the comparison of continuous variables is analyzed by t-test, the classification variables is calculated by chi-square test, the survival analysis between groups is performed by Kaplan-Meier survival curve and log rank test, and the multivariate analysis is done by Cox regression. The risk grade of hypertension complications was judged by the Risk Ratio (RR) of different medication schemes^26, 27^. If RR is ranged from 1.2-1.5, the risk factors are indicated with a weak relationship to complications, while an RR of 1.6-2.9 shows moderate relationship to, and an RR of 3.0 and above illustrates strong relationship to the complications. Besides, the accuracy of risk estimation is judged by the 95% Confidence Interval (CI) of RR. Data analysis was carried out by R software (Version 3.5.2, https://www.r-project.org/).

## 3. Discussion

Artificial intelligence (AI) is developing rapidly in field medic of china, and CDSS is a typical representative. CPR is a core part of CDSS, which stand for a method of recognizing individual patient of cost and effectiveness to help policymakers make more appropriate decisions at the point of medication^28, 29^. Although CPR is important in antihypertensive drugs, the evidence in this respect is still limited in region of China.

To our knowledge, this trial is a first real-world study in China, in which the occurrence of clinical outcomes and medication costs in patients with antihypertensive drugs will be recorded and the relationship between cost-effectiveness of medication and patient characteristics/ therapeutic schedule will be evaluated based on real-world practice. In fact, it has increased its awareness among physicians to provide effective antihypertensive drug scheme according to patient characteristics. Our study will address drug treatment for hypertension in Chinese healthcare facilities, with a particular focus on tertiary hospitals, from a range of real-world perspectives.

As a result, we will construct a two-way study cohort according AHD, and the data of each group include socio-demographic characteristics, disease progression, and clinical outcome^30, 31^. The COX regression model and clinical specialty survey are carried out to identified IF in different study cohorts^21^, and the discriminant model will be used to develop CPR^22^. To evaluate the effect of different medication regimens and patient characteristics on clinical outcomes according to CPR, and the physicians reasonably choose the medication plan according to the evaluation results.

The present analysis is subject to some limitations intrinsic to real world studies, including potential confounding factors^32^ such as polydrug use among users of antihypertensive drugs^33, 34^, adherence to medication^34^, and indication bias^35^ among patients with hypertension. We attempted to minimize these by recruiting the predominant individuals using antihypertensive drugs as the drug of choice within the past six months prior to enrollment to the study.

Importantly, it is noted that the emphasis of the study is a pragmatic approach to clinical decision support for patients who must be accept antihypertensive drugs before long diagnosis of hypertension in real world practice according patient characteristics that are accessible to physicians.

In summary, our research findings hope to provide valuable CPR for physicians and policymakers with respect to treating hypertension according characteristic of individual patients. By developing a predictive method for clinical outcomes and treatment costs of antihypertensive medication, we expect to discriminate those patients who would profit from specific scheme of antihypertensive drugs to minimal incidence probability of costs and complications.

## Data Availability

The datasets used and/or analysis during the current study are available from the corresponding author on reasonable request.

## List of abbreviations

CPR: Clinical Prediction Rules
CDSS: Clinical Decision Support Systems
AHD: Antihypertensive drugs (AHD)
AI: Artificial intelligence
IF: Influencing Factors
EMRC: Electronic Medical Record System
DALY: Disability-Adjusted Life Year

## Declarations

### Ethics approval and consent to participate

Required ethics approvals have been obtained prior to our study from the ethics committee of the First Affiliated Hospital of College of Medicine of Zhejiang University in China, and the committee’s reference number is #2019-1391.

### Ethics and consent to participate

This study conduct by using previously obtained information, and meet four conditions at the same time, including “It is unrealistic or impossible to obtain informed consent”, “The Privacy of subject could be well protected”, “Not more than the minimum risk” and “The right and interests of subject will not be invaded”. Therefore, after careful ethical review by the ethics committee, this study was approved Informed Consent Waiver (Reference number is #2019-1391).

### Competing interests

The authors declare that they have no competing interests.

### Funding

Not applicable.

### Authors’ contributions

DSH and YL had the original idea for this study. WDS and JYW reviewed previous studies. XYL interpreted the patient data regarding antihypertensive drugs. XYL and LL Provide statistical support. DHS registered in chictr and is the guarantor of the protocol. All authors read and approved the final version of the article.

## Acknowledgments

This study is supported by the Zhejiang Province Key Research and Development Program (2019C04006) and Zhejiang Provincial Natural Science Foundation (LQ20G030025) of China. Not applicable.

## Trial registration

This study was registered at www.chictr.org as a primary register of the WHO International Clinical Trials Registry Platform (ICTRP), and the registered number is ChiCTR1900026339.

## Notes

### Competing Interest Statement

The authors have declared no competing interest.

### Clinical Trial

ChiCTR1900026339

